# A one-step real-time RT-PCR assay for simultaneous typing of SARS-CoV-2 mutations associated with the E484K and N501Y spike protein amino-acid substitutions

**DOI:** 10.1101/2021.05.31.21257367

**Authors:** Serafeim C. Chaintoutis, Taxiarchis Chassalevris, George Tsiolas, Sofia Balaska, Ioannis Vlatakis, Evangelia Mouchtaropoulou, Victoria I. Siarkou, Areti Tychala, Dimitris Koutsioulis, Lemonia Skoura, Anagnostis Argiriou, Chrysostomos I. Dovas

## Abstract

The emergence of SARS-CoV-2 mutations resulting in the S protein amino-acid substitutions N501Y and E484K, which have been associated with enhanced transmissibility and immune escape, respectively, necessitates immediate actions, for which their rapid identification is crucial. For the simultaneous typing of both of these mutations of concern (MOCs), a one-step real-time RT-PCR assay employing four locked nucleic acid (LNA) modified TaqMan probes was developed. The assay is highly sensitive with a LOD of 117 copies/reaction, amplification efficiencies >94% and a linear range of over 5 log_10_ copies/reaction. Validation of the assay using known SARS-CoV-2-positive and negative samples from human and animals revealed its ability to correctly identify wild type strains, and strains possessing either one or both targeted amino-acid substitutions, thus comprising a useful pre-screening tool for rapid MOC identification. The basic principles of the methodology for the development of the assay are explained in order to facilitate the rapid design of similar assays able to detect emerging MOCs.

---

Severe acute respiratory syndrome coronavirus 2 (SARS-CoV-2, *Coronaviridae* family) is a novel (+)ssRNA virus responsible for the COVID-19 pandemic (Chan et al., 2020; Dhama et al., 2020; Kim et al., 2020). The viral S (spike) protein mediates receptor binding on target cells and determines host tropism (Lu et al., 2020). Additionally, the S protein is immunogenic and triggers the host specific immune responses, i.e. neutralizing antibodies and T cell-mediated responses; hence, the viral S protein is the main protein used as a target in COVID-19 vaccines (Dai and Gao, 2021). It has been estimated that the SARS-CoV-2 evolves at a rate of approx. 1.1 × 10^−3^ substitutions per site per year (s/s/y) (Martin et al., 2021). Mutations resulting in amino-acid substitutions in the S protein can alter both host cell receptor binding and antigen/antibody binding, with possible effects on infectivity, transmissibility and immune evasion (Di Caro et al., 2021).

Based on analysis of the currently available SARS-CoV-2 genomic sequences via the PANGOLin SARS-CoV-2 lineage assigner interface (Rambaut et al., 2020), several lineages and variants have been assigned. The importance of some SARS-CoV-2 variants (identified as variants of concern, VOCs), has been recognized by the Centers for Disease Control and Prevention (CDC), as they possess attributes that enable increased transmissibility and reduced neutralization activity by convalescent and post-vaccination sera (CDC, 2021). Specifically, the B.1.1.7 lineage (UK variant or VOC 202012/1; 17 amino-acid substitutions) emerged in southeast England in November 2020 and is rapidly spreading towards fixation. It has been indicated that this variant has a higher reproduction number up to 90% higher than pre-existing variants and will lead to large resurgences of COVID-19 cases (Davies et al., 2020). The B.1.351 lineage (South African variant or S.501Y.V2; 17 amino-acid substitutions) was initially reported in South Africa in December 2020. Additionally, the P.1/B.1.1.28.1 lineage (Brazilian variant or 501Y.V3; 17 amino-acid substitutions) was reported in Brazil in January 2021 (Abdool Karim and de Oliveira, 2021). Both of the latter have been also characterized by increased transmissibility.

In all of these VOCs, the N501Y amino-acid substitution (change of asparagine to tyrosine) is present at the position 501 of the S-protein (receptor-binding domain), which confers enhanced affinity for receptor binding (Starr et al., 2020). The two latter VOCs (B.1.351 and P.1) also possess the E484K amino-acid substitution (change from glutamate to lysine), which reduces the neutralization sensitivity to convalescent sera (Wibmer et al., 2021), and thus, possibly affecting the protection conferred by vaccine-derived antibodies, or antibodies produced in previous infections from non-carrying the E484K SARS-CoV-2 strains. These characteristics have a serious impact on the control of the ongoing pandemic, including the need for the modification or the development of novel diagnostic assays, or the currently available vaccines. Most importantly, besides the aforementioned VOCs, both of these substitutions (N501Y and E484K) have been reported to emerge independently in several other SARS-CoV-2 lineages worldwide, such as in lineages AP.1, A.27 for the substitution N501Y, and lineages R.2, N.10 and P.2 for the substitution E484K. As a result, rapid identification of both of the aforementioned mutations of concern (MOCs) is required, so as to immediately take the appropriate public health actions.

The guidelines from the European Centre for Disease Prevention and Control (ECDC) and the World Health Organization (WHO) indicate that SARS-CoV-2 complete genome sequencing, or at least, S gene sequencing (whole or partial), should be used to confirm infection with a specific variant (European Centre for Disease Prevention and Control (ECDC), 2021). Undoubtedly, genome analysis via the next-generation sequencing (NGS) technology comprises the most accurate approach for SARS-CoV-2 molecular characterization. However, the application genome sequencing is labor-intensive, cost-ineffective and may take several hours up to days to be completed, depending on the workflow of the laboratory. According to the guidelines of the aforementioned organizations, alternative methods, such as diagnostic screening PCR-based assays can also be used, for the early detection and prevalence calculation of VOCs (European Centre for Disease Prevention and Control (ECDC), 2021). Many of the currently available PCR-based assays are based on the S gene target failure, associated with the allele “drop-out” phenomenon from targeting deletions alone, or in combination with other traits (Kováčová et al., 2021). As a result, the target of the aforementioned assays is to identify specific VOCs, and not the MOCs associated with the respective amino-acid substitutions possessed by a given variant. On the other hand, SNP-specific assays have been developed, e.g. a melting-curve based assay (Durner et al., 2021), as well as a TaqMan probe-based assay (Sandoval Torrientes et al., 2021), both targeting a single mutation resulting in the amino-acid substitution at the 501 position.

In consideration of above, a novel one-step real-time RT-PCR was developed, for the rapid and accurate simultaneous typing both of the SARS-CoV-2 S gene mutations, associated with the aforementioned substitutions at positions 484 and 501. A 153 bp amplicon of the viral S gene was targeted (positions 22981-23133 on GenBank acc. No. NC_045512), flanking both mutations. Primer Checker tool was used, to search primers against sequences within GISAID’s database of 257,428 high quality SARS-CoV-2 genomes submitted spanning a period from 21/1/2021 up to 19/4/2021. Comparisons for primer SARSpUp2 revealed single and double nucleotide mismatches in 25,130 and 206 genomic sequences, respectively, whereas in the case of primer SARSpDo5, only single mismatches were found in 3,327 sequences (Fig. 1). Primer SARSpUp2 was evaluated *in silico* using the DINAMelt software (Markham and Zuker, 2005), so as to estimate at the annealing temperature of 56 °C, the mole fraction of each oligonucleotide hybridized to the target regions with most common single mismatches found in SARS-CoV-2 genomic sequences (Fig. 1) (Chassalevris et al., 2020). More specifically, for each primer/target variant sequence, the entire equilibrium melting profile was calculated using the DINAMelt application “Hybridization of two different strands of DNA”. The default parameters were used, and the concentration of each unique oligonucleotide (0.2 μM) and salt conditions (50 mM Na^+^, 3.2 mM Mg^2+^) were chosen to correspond to the conditions of the PCR amplification. Estimation of equilibrium melting profiles of the mismatched duplexes indicated that all sequences with single mismatches will be amplified. More specifically, the mole fractions of primer SARSpUp2, hybridized to the targeted variants at 56 °C ranged from 62 to 90%.

**Fig. 1.**
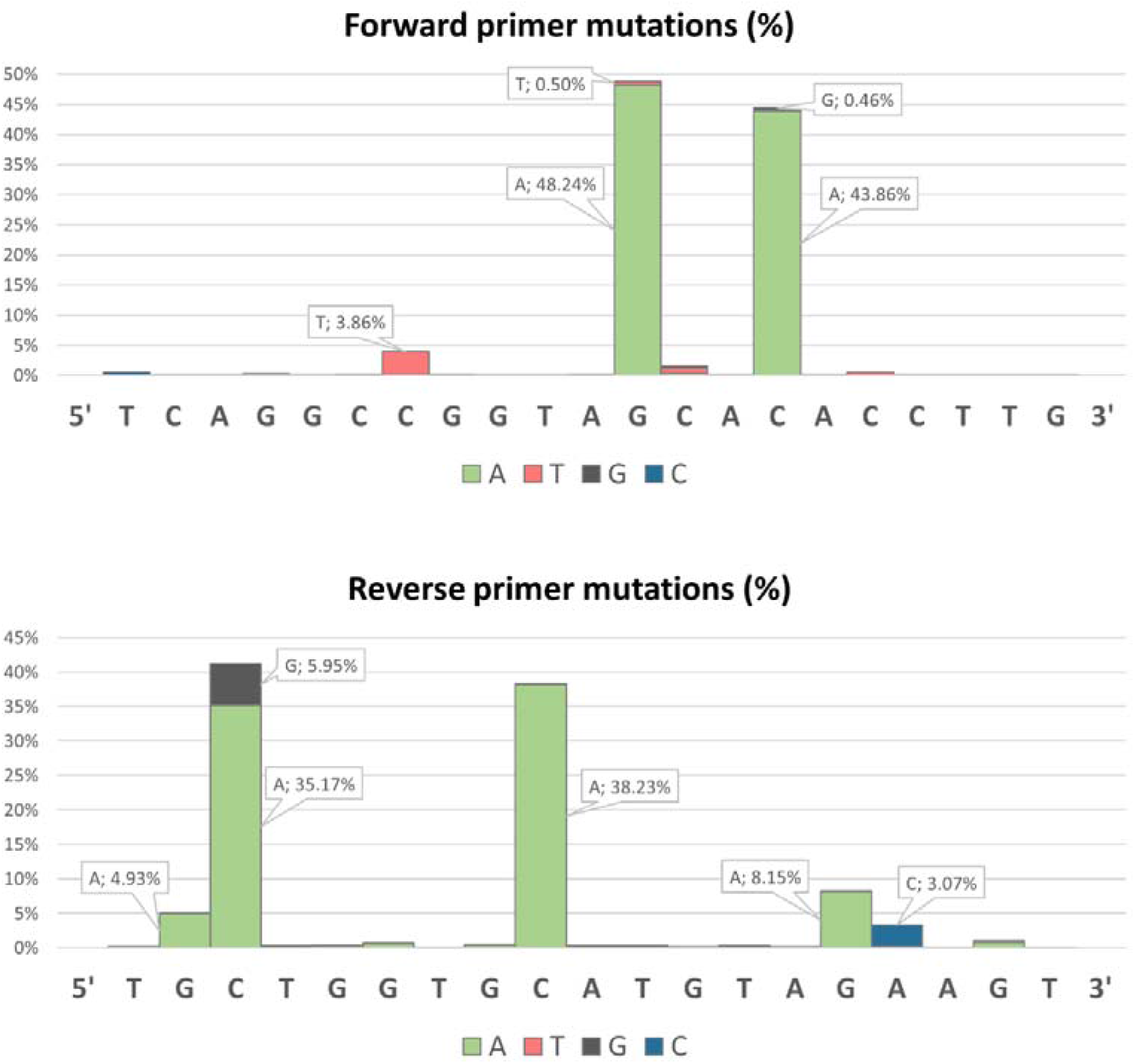
Frequencies of single nucleotide mutations of SARS-CoV-2 homologous S gene sequences spanning a period from 21/1/2021 up to 19/4/2021 compared to primers SARSpUp2 and SARSpDo5.

Four TaqMan probes with locked nucleic acid (LNA) chemistry were designed. Two of them were used for the differentiation between the wild type (WT) and the mutation at position 484, and the other two were used for the same purpose at position 501. Different fluorophores were conjugated at the 5’ end of each TaqMan probe (FAM, HEX, Texas Red and Cy5) to facilitate differentiation of the respective fluorescence signals (Table 1). The effects of LNA modifications on mismatch discrimination were taken into consideration when designing the probes (Owczarzy et al., 2011; You et al., 2006). More specifically a triplet of LNA modifications with the central base of the triplet at the mismatch site were incorporated, whereas modification of a guanine nucleotide or either of its nearest-neighbor bases was avoided in G•T mismatch sites. The probes were short (16 bases) to improve mismatch discrimination. Analysis for the melting temperature (*Tm*), the absence of dimers, and possible hairpin secondary structures was performed using the IDT OligoAnalyzer™ Tool (Integrated DNA Technologies). This tool can also predict stability of LNA-DNA duplexes. Based on our experience on designing similar assays, all probes targeted the same DNA strand and additional LNAs were incorporated in the probes, if needed, so as their *Tm* can be calculated to approximately 63.5 °C. The primer responsible for the extension that degrades the hybridized probes was designed to have a *Tm* 1-2 °C lower, but not less. This procedure for primer design is adopted so as: a) to avoid significant strand extension before the annealing of probes during the PCR cycling and b) to apply annealing temperatures that allow both sufficient mismatch discrimination and sufficient amplification efficiency. The second primer was designed to have a higher *Tm* in order to support a) high amplification efficiency and b) annealing to targets with single mismatches (Table 1, Fig. 1) so as to permit high analytical and diagnostic sensitivity of the assay.

**Table 1.**
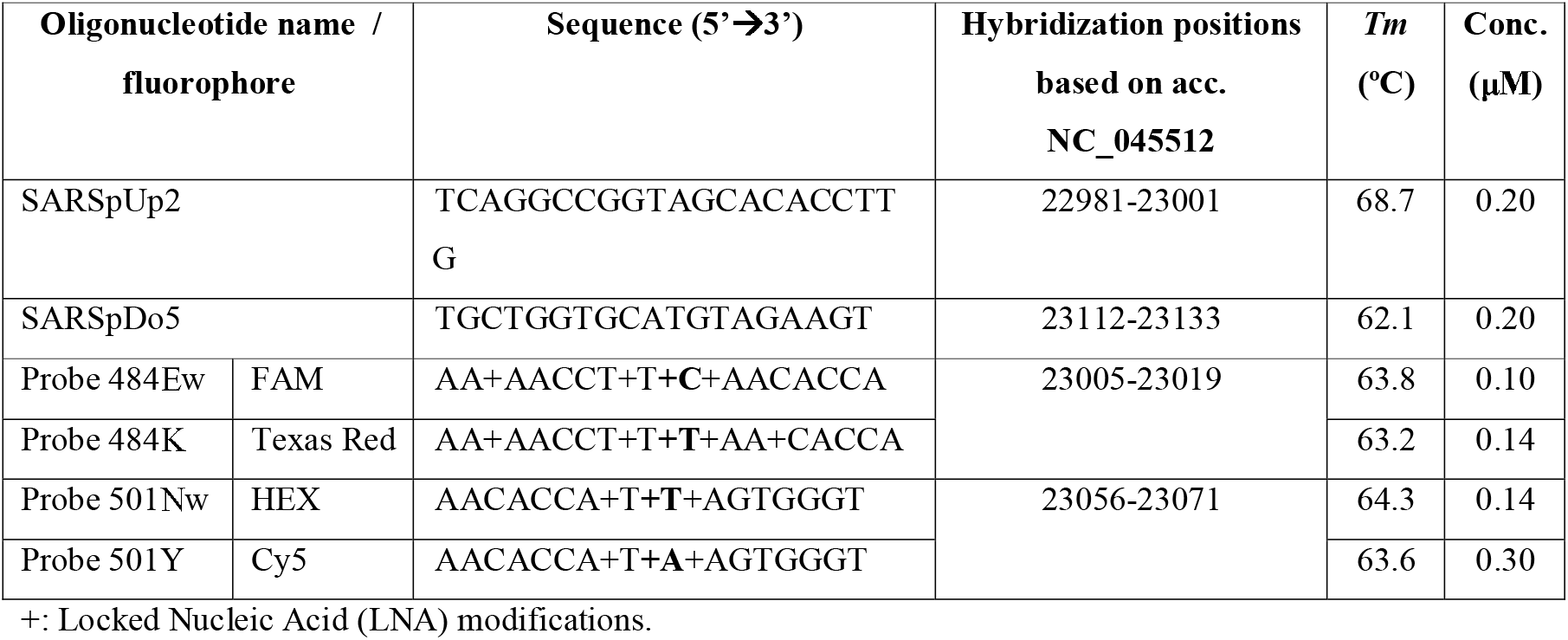
Oligonucleotides (primers and TaqMan probes) used in the developed one-step real-time RT-PCR assay for MOC characterization. All oligonucleotides were synthesized by IDT.

The protocol and composition was developed and optimized using EnzyQuest’s One step RT qPCR kit (Product No.: RN010; EnzyQuest P.C., Heraklion, Greece). Reactions (20 μl) were performed using 4 mM Mg^2+^, the aforementioned oligonucleotides at the concentrations presented in Table 1, and 2 μl of sample DNA extract. The following thermal cycling conditions were applied: 55 °C for 15 min (reverse transcription), 94 °C for 15 min (reverse transcriptase inactivation/Taq polymerase activation), followed by 48 cycles in 2 steps: a) 94 °C for 10 s (denaturation) and b) 56 °C for 40 s (combined annealing/extension). The fluorescence levels were measured at the end of each cycle. The reactions were run on a CFX96 Touch™ Real-Time PCR Detection System (Bio-Rad Laboratories) and the CFX™ Maestro Software (v4.1, Bio-Rad Laboratories) was used for analysis of fluorescence data. Based on the result obtained per each fluorescence channel, the phenotype of the viral strain being typed was characterized as WT i.e. 484E and 501N (FAM/HEX), strain with only the 501Y phenotype (FAM/Cy5, such as UK variant strains), a strain with both MOCs, i.e. 484K and 501Y phenotype (Texas Red/Cy5, such as South African and Brazilian variant strains), or a strain belonging to a different variant with only the E484K amino-acid substitution (Texas Red/HEX) (Fig. 2). Strains of the latter case (only 484K) may belong to recently emerging VOCs associated e.g. with the B.1.526 lineages, which have been reported to rapidly spread in New York (Annavajhala et al., 2021). The B.1.1.318 lineage, which was recently reported by Public Health England and designated as variant under investigation (VUI) is also characterized by only the 484K phenotype in the absence of 501Y substitution (Public Heaalth Ingland, 2021).

**Table 2.** Known SARS-CoV-2 negative (N=26) and positive (N=111) samples tested with the developed assay for validation purposes.

| Sample panel |  | Mutation determination method | No. of samples | Variant | Fluorescence channel |  |  |  |
| --- | --- | --- | --- | --- | --- | --- | --- | --- |
|  |  |  |  |  | FAM | HE X | Texas Red | Cy5 |
| Human | Negative | N/A | 20 | N/A | - | - | - | - |
|  | Positive (A) | Obtained in September 2020, before the emergence of the targeting mutations | 20 | WT | + | + | - | - |
|  |  | ViroBOAR Spike 1.0 RT-PCR Kit (SARS-CoV-2) & Roche LightCycler 480 II instrument | 4 | WT | + | + | - | - |
|  |  |  | 21 | UK (B.1.177) | + | - | - | + |
|  |  |  | 1 | South African (B.1.351) | - | - | + | + |
|  | Positive (B) | NGS (Illumina MiSeq) | 16 | WT (B.1.177) | + | + | - | - |
|  |  |  | 2 | WT (B.1.258) | + | + | - | - |
|  |  |  | 29 | UK (B.1.1.7) | + | - | - | + |
|  |  |  | 12 | South African (B.1.351) | - | - | + | + |
|  |  |  | 1 | Other (B.1.1.318) | - | + | + | - |
| Veterinary | Negative | N/A | 3 cats, 3 minks | N/A | - | - | - | - |
|  | Positive | NGS (Ion Torrent GeneStudio S5) | 2 cats | WT (B.1.1) | + | + | - | - |
|  |  | NGS (Illumina MiSeq) | 3 minks | WT (B.1.1.305) | + | + | - | - |

N/A: not applicable; WT: wild type; NGS: next-generation sequencing

**Fig. 2.**
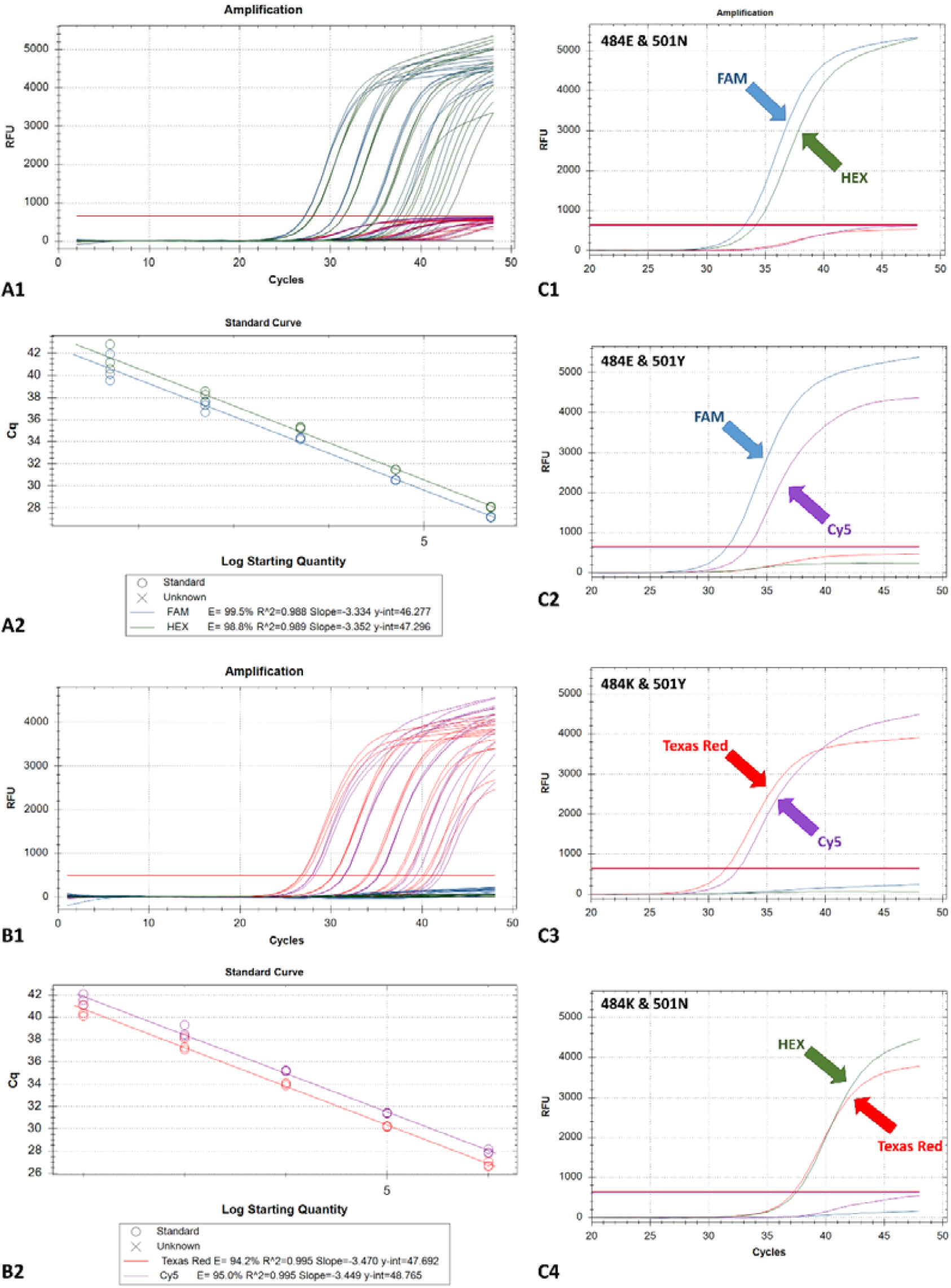
(A1, B1) Amplification plots (fluorescence signals) generated by 2 dilution series of a WT strain (A; FAM/HEX fluorescence channels) and a South African variant strain (B; Texas Red/Cy5 fluorescence channels). Curves represent 10-fold serial dilutions, i.e. from left to right: 5×10^5^ down to 5×10 copies/assay (WT, A1), and from 10^6^ down to 10^2^ copies/assay (South African variant, B1), each tested in 3 replicates. RFU: relative fluorescence units. (A2, B2) The corresponding standard curves. (C1-C4) Interpretation guide from testing representative SARS-CoV-2-positive samples, indicating all possible amino-acid substitution combinations (phenotypes) in the S protein 484 and 501 positions.

For the determination of the analytical characteristics of the developed assay, two positive RNA extracts, i.e. one containing the WT virus (fluorescence channels: FAM/HEX) and one containing the South African variant (fluorescence channels: Texas Red/Cy5) were quantified using the N2 assay by CDC, as previously described (Chaintoutis et al., 2021). Subsequently, two ten-fold dilution series were prepared in a background of a SARS-CoV-2-negative RNA extract from human oropharyngeal swabs, i.e. from 5×10^5^ down to 5×10^1^ copies/assay for the WT, and from 10^6^ down to 10^2^ copies/assay for the South African variant. All prepared dilutions of both dilution series were run in triplicates to determine the amplification efficiency and the linearity of the developed assay against each targeted variant. An amplification efficiency of >94% (i.e. FAM: 99.5%; HEX: 98.8%; Texas Red: 94.2%; Cy5: 95.0%) and a linear range of quantification over 5 log_10_ were observed in all cases (Fig. 2). In order to determine the limit of detection (LOD) both quantified samples were further diluted (between 150 and 12.5 copies/reaction) and tested. Each prepared dilution was tested in octaplicate, and the LOD was calculated with 95% probability of detection, by applying probit regression analysis. Consequently, the LOD for the typing assay was determined at 117 copies/reaction.

The specificity of the developed assay in testing human samples was assessed by testing a panel of SARS-CoV-2-negative human nasopharyngeal swabs (N=20) originating from a university hospital setting. These swabs (as those comprising the positive panel A described below) were obtained for COVID-19 diagnosis, or within the framework of close contact tracing. The SARS-CoV-2-negative status of the samples was based on the results obtained through the application of the Abbott RealTime SARS-CoV-2 assay on the m2000 RealTime System (comprised by the m2000sp sample processor, and the m2000rt thermal cycler; Abbott Laboratories). RNA was extracted using the NucleoSpin RNA Virus kit (Macherey-Nagel) and analysis of the extracts using the developed assay was performed on a CFX96 Touch™ Real-Time PCR Detection System (Bio-Rad Laboratories), revealing the absence of fluorescence in any of the four channels.

The diagnostic performance of the developed assay in MOC identification was validated by testing two panels of SARS-CoV-2-positive human clinical specimens (Panel A: N=46; Panel B: N = 60). Positive panel A samples originated from the same university hospital setting, with those comprising the panel of negative samples. Twenty-six out of the 46 positive samples were collected from February 22^nd^ to April 8^th^, 2021, and were characterized using the ViroBOAR Spike 1.0 RT-PCR Kit (Eurofins Genomics) on a LightCycler 480 II instrument (Roche) and were classified as follows: 21 as UK (B.1.1.7), 1 as South African (B.1.351) and 4 as WT strains (Table 2). Twenty additional WT strains, which were obtained from humans during September 2020, i.e. before the emergence of E484K and N501Y amino-acid substitutions in Greece (National Public Health Organization-Greece (EODY), 2021) were included. The reactions for all samples from the negative panel and the positive panel A were run as described above, on a CFX96 Touch™ Real-Time PCR Detection System (Bio-Rad Laboratories) and all of them were correctly identified.

Samples comprising positive panel B were obtained from humans in Northern Greece (Thessaloniki, and nearby locations) for SARS-CoV-2 diagnosis and within the framework of the National Flagship Action “Greece vs Corona”, except for one sample which was obtained in Athens. RNA extraction was performed using the MagMAX™ Viral/Pathogen Nucleic Acid Isolation kit (ThermoFisher Scientific) on the KingFisher™ Flex instrument (ThermoFisher Scientific). The viral strains from Northern Greece were characterized by whole genome sequencing using the NGS technology (Illumina MiSeq) and were classified as follows: 29 as UK (B.1.1.7), 12 as South African (B.1.351) and 18 as WT strains (B.1.285 & B.1.177). The viral strain obtained from Athens was characterized as variant under investigation (B.1.1.318) as only the 484K phenotype was present. Testing of these RNA extracts using the developed assay was performed as described above, on a Rotor-Gene Q 5plex Platform (Qiagen). The channels used for fluorescence acquisition were Green, Yellow, Orange and Red, for HEX, FAM, Texas Red and Cy5 fluorophores, respectively. The Rotor-Gene Q Application Software was used for data analysis, through the function “Allelic Discrimination”. Overall, the developed assay was able to accurately identify both MOCs and discriminate between the different tested variants (Table 2). Low levels of non-specific fluorescence occurred in the Texas Red channel, without affecting the analysis process and the obtained results.

Additionally, given that some animal species are susceptible to SARS-CoV-2, a veterinary collection of samples was tested via the developed method. Specifically, a panel of negative samples originating from cats and minks (N = 3 for each species) was tested, revealing the absence of fluorescence in any of the four channels. SARS-CoV-2-positive oropharyngeal swabs from cats which were investigated in a previous work of our team (Chaintoutis et al., 2021) were also tested. SARS-CoV-2-positive oropharyngeal samples from minks were also tested. Specifically, 3 animals infected by a WT strain (B.1.1.305) originating from a heavily infected farm were tested. All animal samples were correctly identified regarding the detection of the relevant mutations (Table 2).

In conclusion, the typing real-time RT-PCR assay developed herein is able to accurately identify the mutations associated with the E484K and N501Y phenotypes of SARS-CoV-2 S protein, considered as MOCs. The assay is simple to perform, rapid, sensitive and can be applied in a variety of specimens from human and animals. Our methodology of primer and probe design can support both high sensitivity of detection and sufficient mismatch discrimination and can facilitate the rapid design of similar assays able to detect emerging MOCs. The results suggest that the developed system is useful as a fast pre-screening tool for the selection of VOCs and other variants of interest (VOIs) for subsequent characterization via NGS analysis, or for MOCs prevalence calculation within the framework of epidemiological investigations.

## Data Availability

N/A

## Acknowledgments

We thank Dr. Anastasia Chatzidimitriou for providing testing samples.

## CRediT authorship contribution statement

**Serafeim C. Chaintoutis:** Formal analysis, Investigation, Visualization, Writing – original draft. **Taxiarchis Chassalevris:** Investigation, Methodology. **George Tsiolas:** Investigation. **Sofia Balaska:** Investigation. **Dimitris Koutsioulis:** Investigation, Resources. **Evangelia Mouchtaropoulou:** Investigation. **Victoria I. Siarkou:** Investigation, Resources, Writing – review & editing. **Areti Tychala:** Investigation. **Ioannis Vlatakis:** Investigation, Resources. **Lemonia Skoura:** Resources, Validation, Writing – review & editing. **Anagnostis Argiriou:** Resources, Validation, Writing – original draft, Writing – review & editing. **Chrysostomos I. Dovas:** Conceptualization, Methodology, Supervision, Writing – review & editing.

## References

Abdool Karim, S.S., de Oliveira, T., 2021. New SARS-CoV-2 Variants — Clinical, Public Health, and Vaccine Implications. N. Engl. J. Med. NEJMc2100362. https://doi.org/10.1056/NEJMc2100362

Annavajhala, M.K., Mohri, H., Zucker, J.E., Sheng, Z., Wang, P., Gomez-Simmonds, A., Ho, D.D., Uhlemann, A.-C., 2021. A Novel SARS-CoV-2 Variant of Concern, B.1.526, Identified in New York. medRxiv Prepr. Serv. Heal. Sci. https://doi.org/10.1101/2021.02.23.21252259

CDC, 2021. SARS-CoV-2 Variants of Concern [WWW Document]. URL https://www.cdc.gov/coronavirus/2019-ncov/cases-updates/variant-surveillance/variant-info.html (accessed 4.8.21).

Chaintoutis, S.C., Siarkou, V.I., Mylonakis, M.E., Kazakos, G.M., Skeva, P., Bampali, M., Dimitriou, M., Dovrolis, N., Polizopoulou, Z.S., Karakasiliotis, I., Dovas, C.I., 2021. Limited crossLJspecies transmission and absence of mutations associated with SARS□CoV□2 adaptation in cats: A case study of infection in a small household setting. Transbound. Emerg. Dis. tbed.14132. https://doi.org/10.1111/tbed.14132

Chan, J.F.W., Kok, K.H., Zhu, Z., Chu, H., To, K.K.W., Yuan, S., Yuen, K.Y., 2020. Genomic characterization of the 2019 novel human-pathogenic coronavirus isolated from a patient with atypical pneumonia after visiting Wuhan. Emerg. Microbes Infect. 9, 221–236. https://doi.org/10.1080/22221751.2020.1719902

Chassalevris, T., Chaintoutis, S.C., Apostolidi, E.D., Giadinis, N.D., Vlemmas, I., Brellou, G.D., Dovas, C.I., 2020. A highly sensitive semi-nested real-time PCR utilizing oligospermine-conjugated degenerate primers for the detection of diverse strains of small ruminant lentiviruses. Mol. Cell. Probes 101528. https://doi.org/10.1016/j.mcp.2020.101528

Dai, L., Gao, G.F., 2021. Viral targets for vaccines against COVID-19. Nat. Rev. Immunol. https://doi.org/10.1038/s41577-020-00480-0

Davies, N.G., Barnard, R.C., Jarvis, C.I., Kucharski, A.J., Munday, J., Pearson, C.A.B., Russell, T.W., Tully, D.C., Abbott, S., Gimma, A., Waites, W., Wong, K.L.M., van Zandvoort, K., Eggo, R.M., Funk, S., Jit, M., Atkins, K.E., Edmunds, W.J., Houben, R., Meakin, S.R., Quilty, B.J., Liu, Y., Flasche, S., Lei, J., Sun, F.Y., Krauer, F., Lowe, R., Bosse, N.I., Nightingale, E.S., Sherratt, K., Abbas, K., O’Reilly, K., Gibbs, H.P., Villabona-Arenas, C.J., Waterlow, N.R., Medley, G., Brady, O., Williams, J., Rosello, A., Klepac, P., Koltai, M., Sandmann, F.G., Foss, A.M., Jafari, Y., Prem, K., Chan, Y.W.D., Hellewell, J., Procter, S.R., Jombart, T., Knight, G.M., Endo, A., Quaife, M., Showering, A., Clifford, S., 2020. Estimated transmissibility and severity of novel SARS-CoV-2 Variant of Concern 202012/01 in England. medRxiv. https://doi.org/10.1101/2020.12.24.20248822

Dhama, K., Patel, S.K., Sharun, K., Pathak, M., Tiwari, R., Yatoo, M.I., Malik, Y.S., Sah, R., Rabaan, A.A., Panwar, P.K., Singh, K.P., Michalak, I., Chaicumpa, W., Martinez-Pulgarin, D.F., Bonilla-Aldana, D.K., Rodriguez-Morales, A.J., 2020. SARS-CoV-2 jumping the species barrier: Zoonotic lessons from SARS, MERS and recent advances to combat this pandemic virus. Travel Med. Infect. Dis. https://doi.org/10.1016/j.tmaid.2020.101830

Di Caro, A., Cunha, F., Petrosillo, N., Beeching, N., Ergonul, O., Petersen, E., Koopmans, M., 2021. SARS-CoV-2 escape mutants and protective immunity from natural infections or immunizations. Clin. Microbiol. Infect. https://doi.org/10.1016/j.cmi.2021.03.011

Durner, J., Burggraf, S., Czibere, L., Tehrani, A., Watts, D.C., Becker, M., 2021. Fast and cost-effective screening for SARS-CoV-2 variants in a routine diagnostic setting. Dent. Mater. 37, e95–e97. https://doi.org/10.1016/j.dental.2021.01.015

European Centre for Disease Prevention and Control (ECDC), 2021. Methods for the detection and identification of SARS-CoV-2 variants.

Kim, D., Lee, J.Y., Yang, J.S., Kim, J.W., Kim, V.N., Chang, H., 2020. The Architecture of SARS-CoV-2 Transcriptome. Cell 181, 914-921.e10. https://doi.org/10.1016/j.cell.2020.04.011

Kováčová, V., Boršová, K., Paul, E.D., Radvánszka, M., Hajdu, R., čabanová, V., Sláviková, M., Ličková, M., Lukáčiková, Ľ., Belák, A., Roussier, L., Kostičová, M., Líšková, A., Maďarová, L., štefkovičová, M., Reizigová, L., Nováková, E., Sabaka, P., Koščálová, A., Brejová, B., Staroňová, E., Mišík M., Vinař, T., Nosek, J., čekan, P., Klempa, B., 2021. Surveillance of SARS-CoV-2 lineage B.1.1.7 in Slovakia using a novel, multiplexed RT-qPCR assay. medRxiv 2021.02.09.21251168. https://doi.org/10.1101/2021.02.09.21251168

Lu, R., Zhao, X., Li, J., Niu, P., Yang, B., Wu, H., Wang, W., Song, H., Huang, B., Zhu, N., Bi, Y., Ma, X., Zhan, F., Wang, L., Hu, T., Zhou, H., Hu, Z., Zhou, W., Zhao, L., Chen, J., Meng, Y., Wang, J., Lin, Y., Yuan, J., Xie, Z., Ma, J., Liu, W.J., Wang, D., Xu, W., Holmes, E.C., Gao, G.F., Wu, G., Chen, W., Shi, W., Tan, W., 2020. Genomic characterisation and epidemiology of 2019 novel coronavirus: implications for virus origins and receptor binding. Lancet 395, 565– 574. https://doi.org/10.1016/S0140-6736(20)30251-8

Markham, N.R., Zuker, M., 2005. DINAMelt web server for nucleic acid melting prediction. Nucleic Acids Res. 33, W577–W581. https://doi.org/10.1093/nar/gki591

Martin, M.A., VanInsberghe, D., Koelle, K., 2021. Insights from SARS-CoV-2 sequences. Science (80-.). https://doi.org/10.1126/science.abf3995

National Public Health Organization-Greece (EODY), 2021. Information on the results of the Genomic Surveillance Network for SARS-CoV-2 mutations [in greek] [WWW Document]. URL https://eody.gov.gr/enimerosi-schetika-me-ta-apotelesmata-diktyoy-gonidiomatikis-epitirisis-gia-tis-metallaxeis-toy-sars-cov-2/#main-container (accessed 4.28.21).

Owczarzy, R., You, Y., Groth, C.L., Tataurov, A. V., 2011. Stability and mismatch discrimination of locked nucleic acid-DNA duplexes. Biochemistry 50, 9352–9367. https://doi.org/10.1021/bi200904e

Public Heaalth Ingland, 2021. SARS-CoV-2 variants of concern and variants under investigation in England.

Rambaut, A., Holmes, E.C., O’Toole, A., Hill, V., McCrone, J.T., Ruis, C., du Plessis, L., Pybus, O.G., 2020. A dynamic nomenclature proposal for SARS-CoV-2 lineages to assist genomic epidemiology. Nat. Microbiol. 5, 1403–1407.

Sandoval Torrientes, M., Castelló Abietar, C., Boga Riveiro, J., Álvarez-Argüelles, M.E., Rojo-Alba, S., Abreu Salinas, F., Costales González, I., Pérez Martínez, Z., Martín Rodríguez, G., Gómez de Oña, J., Coto García, E., Melón García, S., 2021. A novel single nucleotide polymorphism assay for the detection of N501Y SARS-CoV-2 variants. J. Virol. Methods 114143. https://doi.org/10.1016/j.jviromet.2021.114143

Starr, T.N., Greaney, A.J., Hilton, S.K., Ellis, D., Crawford, K.H.D., Dingens, A.S., Navarro, M.J., Bowen, J.E., Tortorici, M.A., Walls, A.C., King, N.P., Veesler, D., Bloom, J.D., 2020. Deep Mutational Scanning of SARS-CoV-2 Receptor Binding Domain Reveals Constraints on Folding and ACE2 Binding. Cell 182, 1295-1310.e20. https://doi.org/10.1016/j.cell.2020.08.012

Wibmer, C.K., Ayres, F., Hermanus, T., Madzivhandila, M., Kgagudi, P., Oosthuysen, B., Lambson, B.E., de Oliveira, T., Vermeulen, M., van der Berg, K., Rossouw, T., Boswell, M., Ueckermann, V., Meiring, S., von Gottberg, A., Cohen, C., Morris, L., Bhiman, J.N., Moore, P.L., 2021. SARS-CoV-2 501Y.V2 escapes neutralization by South African COVID-19 donor plasma. Nat. Med. 1–4. https://doi.org/10.1038/s41591-021-01285-x

You, Y., Moreira, B.G., Behlke, M.A., Owczarzy, R., 2006. Design of LNA probes that improve mismatch discrimination. Nucleic Acids Res. 34. https://doi.org/10.1093/nar/gkl175

